# Feasibility of time-restricted eating during pregnancy and effect on glycemic control in people with increased risk of gestational diabetes – a randomized controlled trial

**DOI:** 10.1101/2024.05.06.24306931

**Authors:** HS Skarstad, KL Haganes, MAJ Sujan, TM Gellein, MK Johansen, KÅ Salvesen, JA Hawley, T. Moholdt

## Abstract

Time-restricted eating (TRE) is a nutritional intervention that confines the daily time-window for energy intake. TRE reduces fasting glucose concentrations in non-pregnant individuals, but whether this eating protocol is feasible and effective for glycemic control in pregnancy is unknown. The aim of this randomized controlled trial was to investigate the feasibility and effect of a 5-week TRE intervention among pregnant individuals at risk of gestational diabetes mellitus (GDM), compared with a usual-care control group. Participants underwent 2-h oral glucose tolerance tests and estimation of body composition, before and after the intervention. Interstitial glucose levels were continuously measured, and adherence rates and ratings of hunger were recorded daily. Thirty of 32 participants completed the trial. Participants allocated to TRE reduced their daily eating window from 12.3 (SD 1.3) to 9.9 (SD 1.0) h, but TRE did not affect glycemic measures, blood pressure, or body composition, compared with the control group. TRE increased hunger levels in the evening, but not in the morning, and induced only small changes in dietary intake. A 5-week TRE intervention was feasible for pregnant individuals with increased risk of GDM but had no effect on cardiometabolic outcomes.

## Introduction

The prevalence of diabetes is increasing in parallel with the obesity pandemic, with gestational diabetes mellitus (GDM) estimated to occur in up to 14% of all pregnancies.^1^GDM is the development of glucose intolerance with onset or first recognition during pregnancy, brought on by an underlying chronic insulin resistance due to beta-cell dysfunction.^2,3^ Important risk factors for GDM include advanced maternal age, a body mass index (BMI) ≥ 25 kg/m^2^, previous GDM or a family history with diabetes, delivery of a macrosomic child, and non-Caucasian ethnicity.^4,5^ Glucose intolerance during pregnancy increases the risk of adverse pregnancy outcomes, such as pre-eclampsia, pre-term birth, macrosomia, caesarean delivery, and birth injury.^2,6,7^ Furthermore, GDM increases the risk of developing diabetes and cardiovascular disease later in life for both the mother and the offspring.^2,7,8^

Lifestyle interventions, including nutritional therapy, is regarded as the primary strategy for managing GDM.^5,9^ However, there is no consensus on which diet is best for achieving optimal glycemic control in pregnancy.^5,10^ Time-restricted eating (TRE) is a dietary strategy in which the time-window for energy intake each day is restricted, typically from 6-10 h/day. TRE has shown to have positive effects on glucose regulation in people with overweight/obesity^11,12^ and to improve body composition.^13^ As such, TRE has emerged as a potentially beneficial intervention for individuals at risk of, or diagnosed with GDM.^14^ However, we are unaware of published research on the feasibility or effect of TRE in pregnant individuals. The primary aim of this randomized controlled trial (RCT) was to examine the feasibility of TRE in pregnant individuals with at least one risk factor for developing GDM. Secondary outcomes included the effect of TRE on markers of metabolic health and glycemic control. We hypothesized that it would be feasible for pregnant individuals to adhere to TRE during the second or third trimester, and that TRE would improve glycemic control.

## Results

### Participants

Thirty-two participants undertook baseline assessments and were randomized to either TRE (*n* = 15) or a control group (*n* = 17) (Figure 1). The first participant was recruited 18.01.2019 and the last date of follow-up was 13.03.2023, with a halt in inclusion of participants from March to September 2020 during the Covid pandemic. We stopped the inclusion of participants when we had reached our pre-specified number of participants. Two individuals withdrew their consent to participate in the study after baseline testing and randomization. All participants had at least one risk factor for GDM according to the Norwegian recommendations for screening of GDM, with some having more than one risk factor. Most of the included participants (*n* = 28) had a pre-pregnancy BMI ≥ 25 kg/m^2^. Nineteen were expecting their first child at an age ≥ 25 years, two were of Asian or African ethnicity, three had first-degree relatives with diabetes, two had previously given birth to a child with birthweight > 4.5 kg, and one had been diagnosed with GDM in a previous pregnancy. Table 1 shows an overview of baseline characteristics according to group.

**Figure 1.**
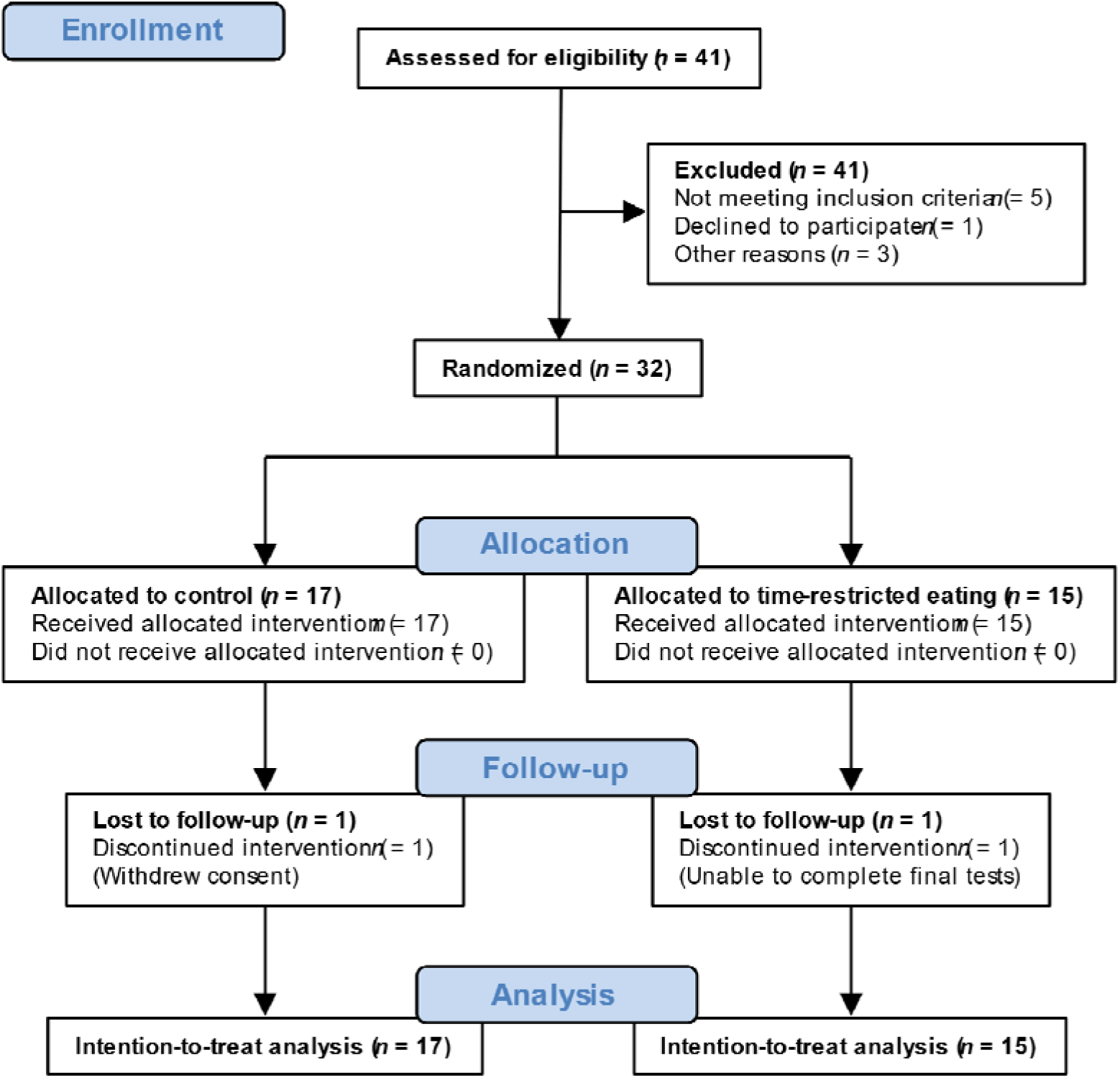
Flow diagram of participants in the study.

**Table 1.** Baseline characteristics of participants, according to group allocation. Data are means with standard deviation (SD).

|  | *n* | Control (*n* = 17) | *n* | Time-restricted eating (*n* = 15) |
| --- | --- | --- | --- | --- |
| Age, years | 17 | 30.1 (2.9) | 15 | 32.2 (3.8) |
| Gestational week | 17 | 19.8 (5.2) | 15 | 18.6 (6.7) |
| Parity | 17 | 0.5 (0.7) | 15 | 0.7 (0.8) |
| Height, cm | 17 | 169 (7) | 15 | 163 (9) |
| Weight, kg | 17 | 81.3 (15.2) | 15 | 79.6 (16.4) |
| Body mass index, kg/m<sup>2</sup> | 17 | 28.5 (5.3) | 15 | 29.9 (5.5) |
| Muscle mass, kg | 17 | 28.5 (4.1) | 15 | 26.9 (4.2) |
| Fat mass, kg | 17 | 29.7 (11.0) | 15 | 30.8 (11.7) |
| Fat percentage | 17 | 35.6 (7.5) | 15 | 37.8 (7.3) |
| Visceral fat area, cm<sup>2</sup> | 17 | 140 (54) | 15 | 147 (60) |
| Systolic BP, mmHg | 16 | 115 (7) | 15 | 113 (10) |
| Diastolic BP, mmHg | 16 | 76 (5) | 15 | 74 (6) |
| Heart rate, beats/min | 16 | 77 (12) | 15 | 80 (10) |
| Fasting glucose, mmol/L | 17 | 4.5 (0.4) | 15 | 4.5 (0.3) |
| 120-min glucose, mmol/L | 17 | 4.9 (1.3) | 15 | 5.3 (1.8) |
| Fasting insulin, µIU/mL | 17 | 14.2 (5.4) | 15 | 16.5 (8.7) |
| 120-min insulin, µIU/mL | 17 | 42.0 (29.0) | 15 | 69.8 (97.4) |
| HbA1c, mmol/mol | 17 | 29.8 (3.1) | 14 | 31.0 (3.4) |
| HOMA2-IR | 17 | 1.7 (0.6) | 15 | 2.0 (1.0) |
| Cholesterol, mmol/L | 17 | 5.4 (1.0) | 13 | 5.7 (0.9) |
| Triglycerides, mmol/L | 17 | 1.5 (0.4) | 13 | 1.5 (0.5) |
| HDL, mmol/L | 17 | 2.0 (0.4) | 13 | 2.0 (0.4) |
| LDL, mmol/L | 17 | 3.4 (0.8) | 13 | 3.7 (0.8) |

BP = Blood pressure, HDL = High-density lipoprotein cholesterol, HOMA2-IR = Homeostatic model assessment of insulin resistance, LDL = Low-density lipoprotein cholesterol.

### Time-restricted eating was feasible in pregnancy and affected feelings of hunger

The participants allocated to TRE reduced their eating window from 12.0 h (SD 2.2) at baseline to 9.9 h (SD 1.6) during the 5-week intervention period, with no change in eating window in the control group (12.5 h (SD 1.9) at baseline and 13.1 h (SD 1.7) during the intervention period) (Figure 2). During the baseline week, the participants in the TRE group consumed their first meal at 09:17 h (SD 2.0) and their last meal at 21:01 h (SD 1.1), whereas the average times for the first and last meal during the intervention were 08:43 h (SD 1.9) and 18:50 h (SD 1.7), respectively (Figure 3). Individuals in the TRE group adhered to the ≤ 10-h eating window on 4.7 (SD 0.4) days/week during the intervention period, giving an adherence rate of 67% (SD 6%).

**Figure 2.**
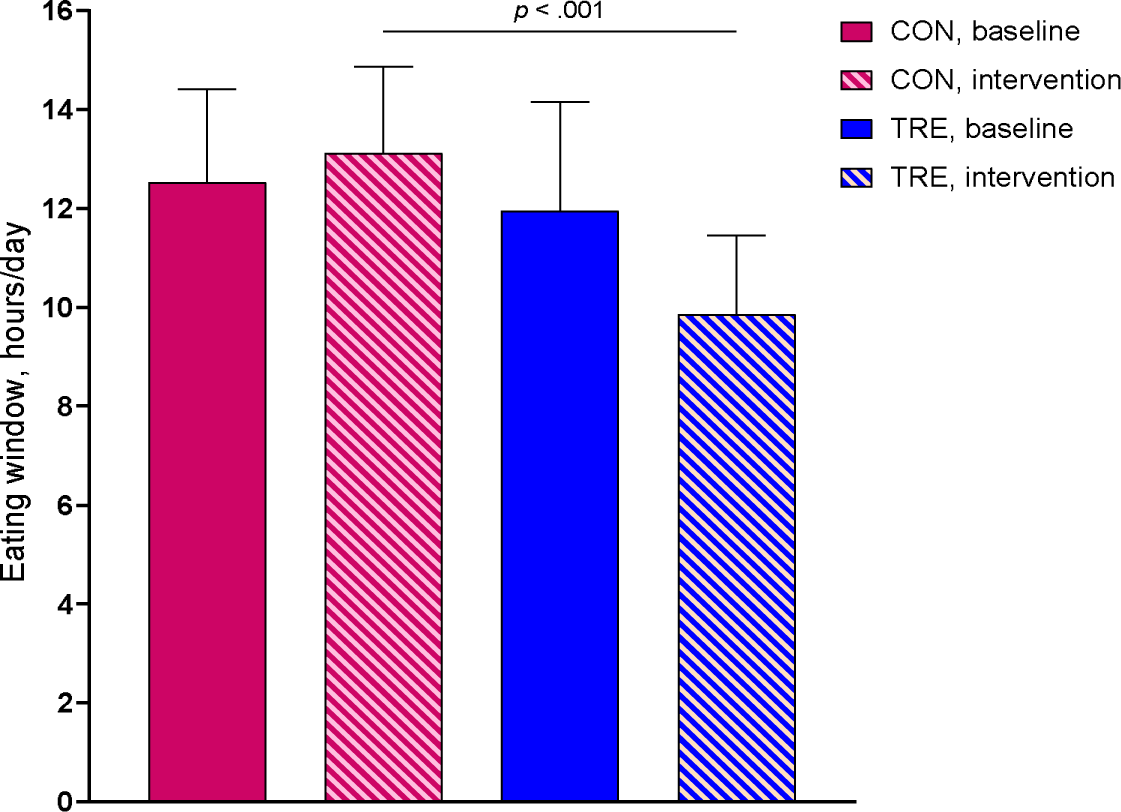
Mean eating window duration. Observed values at baseline and during the 5-week intervention period according to group. Descriptive statistics with standard deviation for the intention-to-treat population. p – value was computed using linear mixed model, comparing the time-restricted eating (TRE) group with the control group (CON).

**Figure 3.**
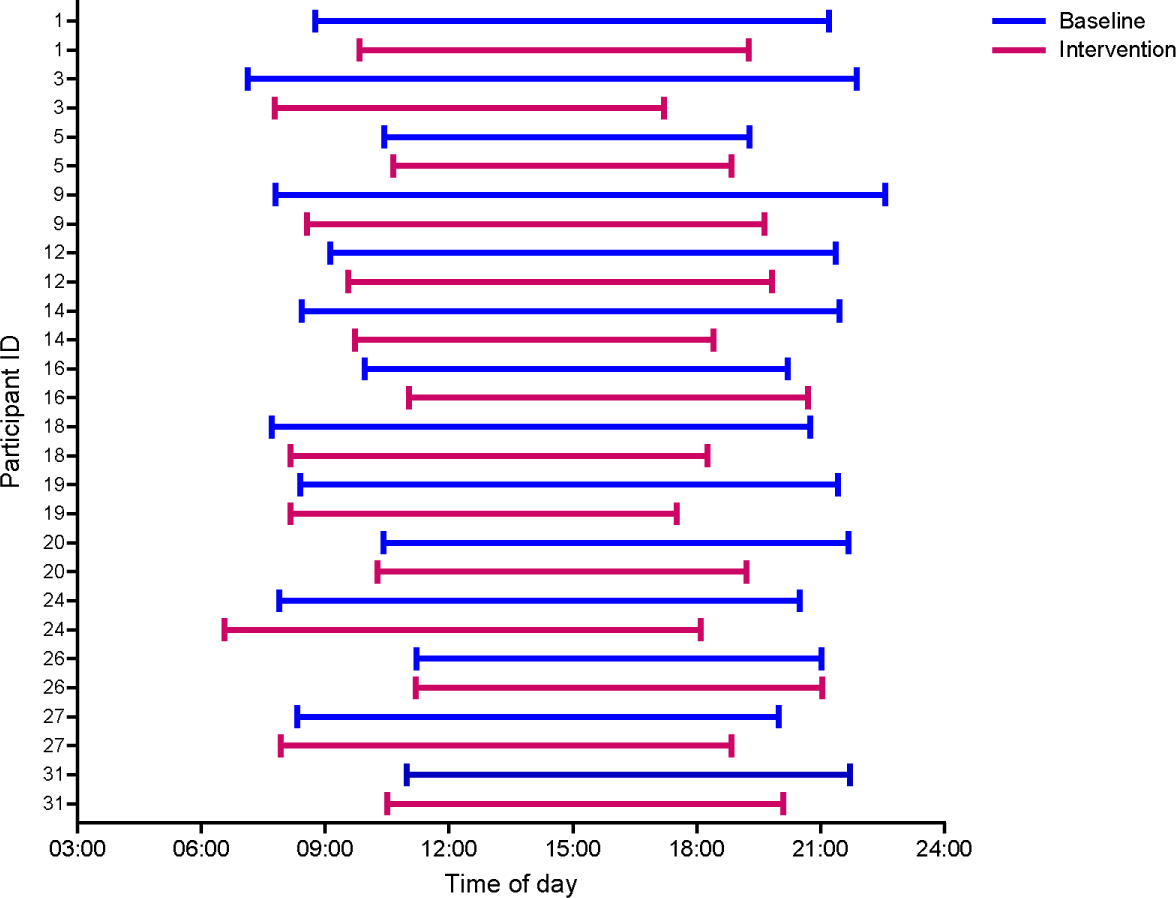
Individual changes in eating window for participants allocated to time-restricted eating. Lines show the average self-reported window for energy intake per day for each participant in the intervention group. The graph is based on observed values from participant handbooks.

There were no effects of the TRE intervention on the participants’ ratings of hunger in the morning (Figure 4, Supplementary Table 1). In the evening during the second week of the intervention, the participants in TRE reported increased hunger levels, desire to eat, and prospective intake of energy, and decreased satiety, compared with CON (Figure 5, Supplementary Table 1). Evening hunger was still higher and evening satiety lower in the TRE group in the last week of the intervention, compared with CON, but the desire to eat in the evening was not.

**Figure 4.**
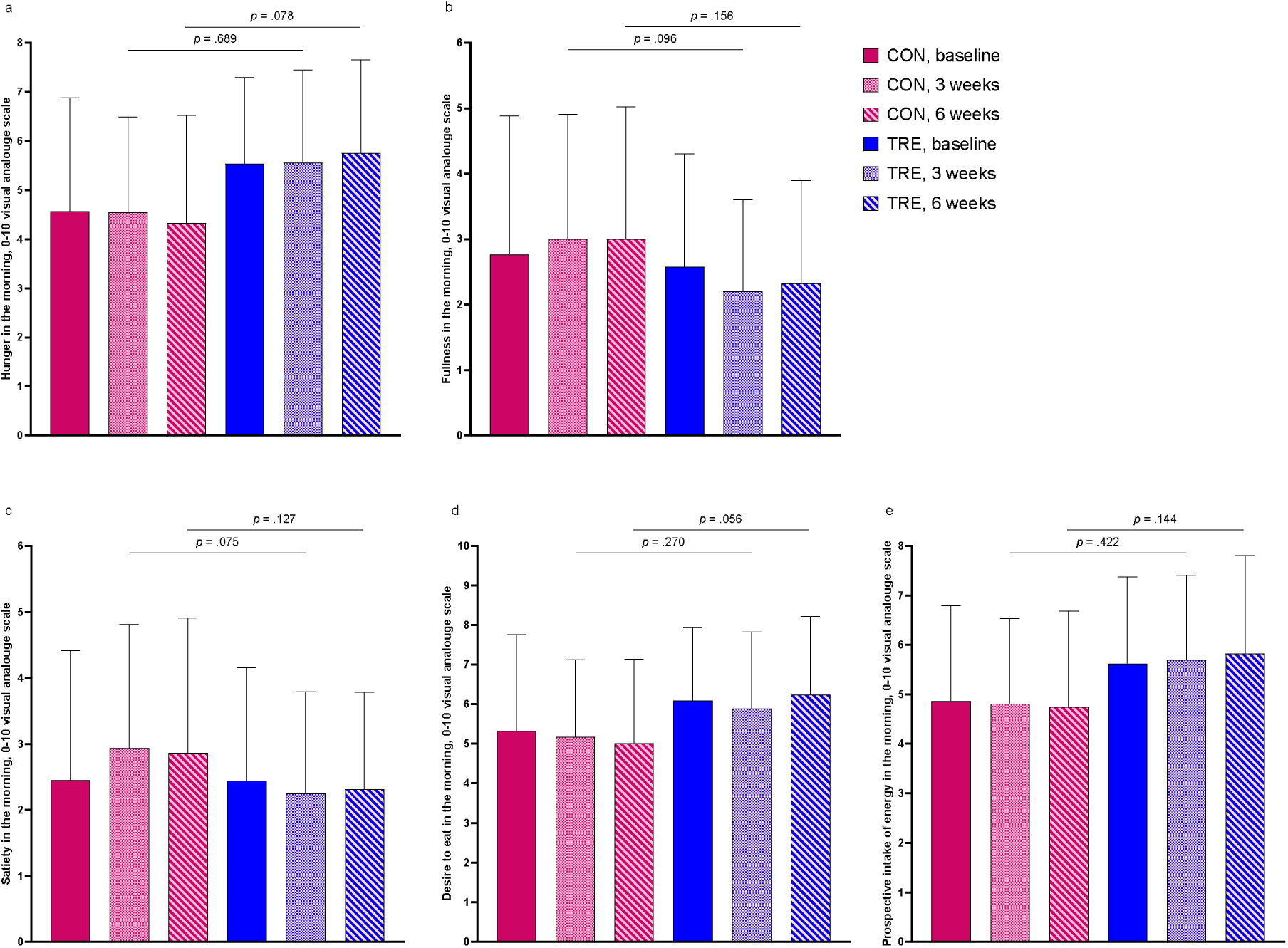
Self-reported appetite in the morning. Feelings of a) hunger, b) fullness, c) satiety, d) desire to eat, and e) prospective intake of energy, as indicated on a 0-10 visual analogue scale in the morning before the first energy intake. The data are observed mean scores at baseline, in the second intervention week (3 weeks) and in the last intervention week (6 weeks). Bars show averages and error bars show standard deviations. p – values are for between-group comparisons using linear mixed models. CON = Control group, TRE = Time-restricted eating.

**Figure 5.**
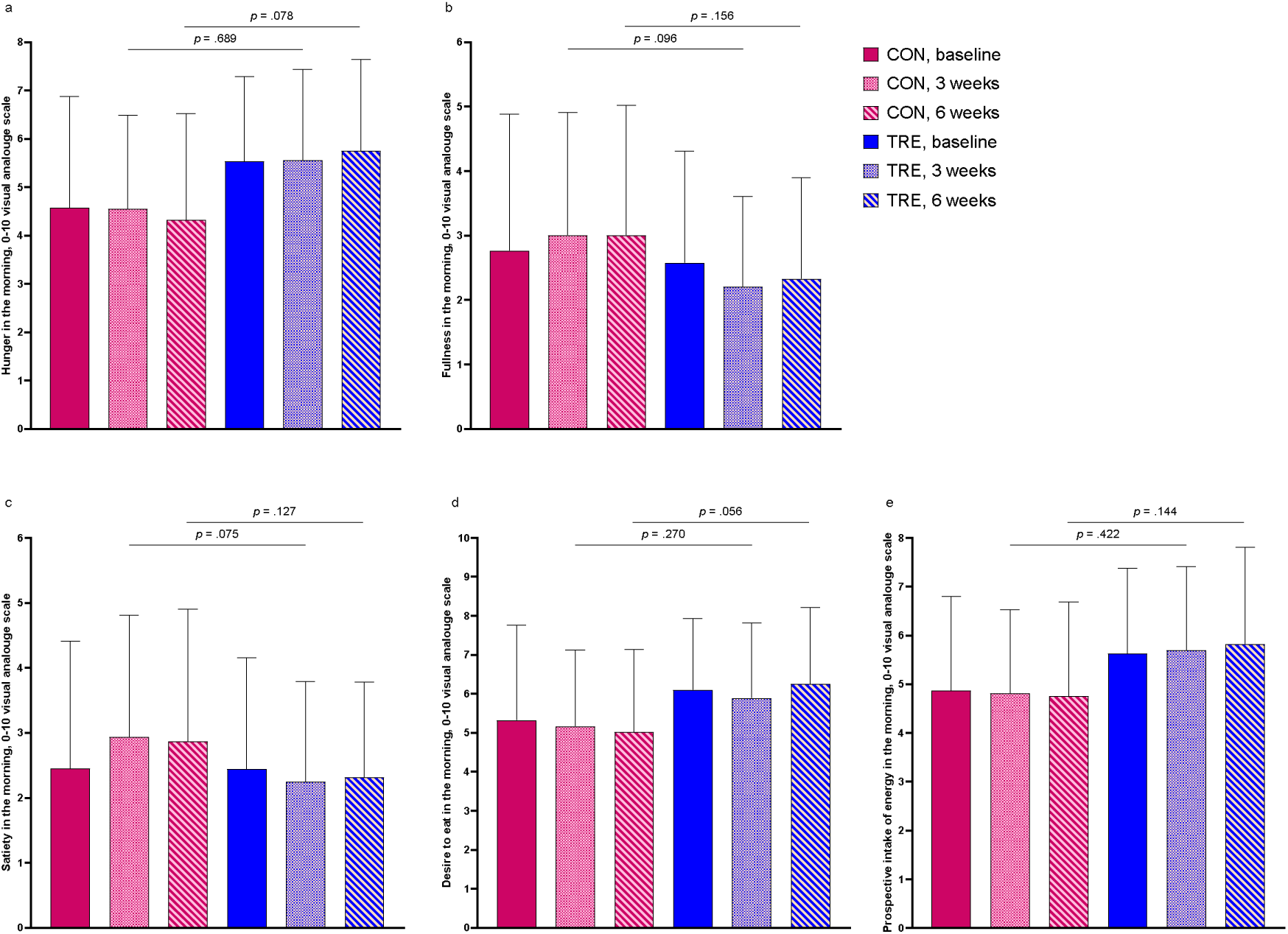
Self-reported appetite in the evening. Feelings of a) hunger, b) fullness, c) satiety, d) desire to eat, and e) prospective intake of energy, as indicated on a 0-10 visual analogue scale in the evening before going to bed. The data are observed mean scores at baseline, in the second week of the intervention period (3 weeks) and in the last week of the intervention period (6 weeks). Graphs show averages and error bars show standard deviations. p – values are for between-group comparisons using linear mixed models. CON = Control group, TRE = Time-restricted eating.

### Time-restricted eating had no effect on secondary metabolic outcomes

At baseline, none of the participants fulfilled the Norwegian criteria for GDM: fasting glucose between 5.3-6.9 mmol/L and/or 120-min glucose between 9.0-11.0 mmol/L after a 75-g oral glucose tolerance test (OGTT).^15^ At the testing after the intervention period, one participant in CON had fasting glucose of 5.7 mmol/L, with all remaining participants being below the threshold for GDM. TRE had no effect on any of the glycemic or metabolic outcomes (Table 2), nor on 24-h glucose area under the curve (AUC), day-time glucose AUC or night-time glucose AUC (Figure 6, Supplementary Table 2). The TRE intervention had no effect on systolic or diastolic blood pressure (Table 2).

**Figure 6.**
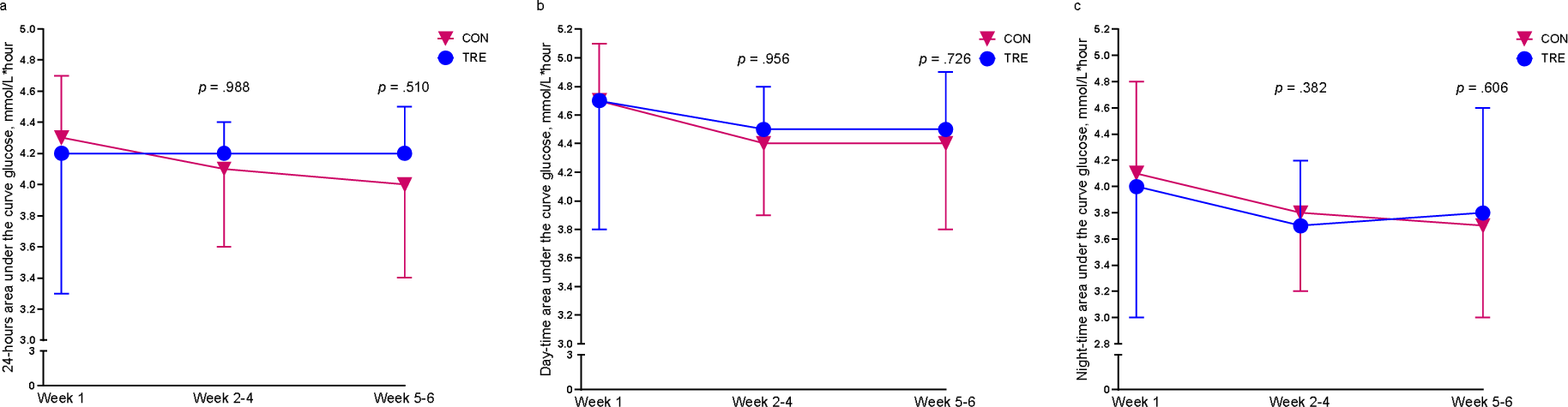
Area under curve (AUC) glucose. a) 24-hours AUC, b) day-time AUC, and c) night-time AUC. Data are estimated from continuous glucose monitoring using Glyculator 3.0. Symbols show averages and error bars show standard deviations for the control group (CON) and the time-restricted eating group (TRE) in the baseline week (Week 1), the first three weeks of the intervention period (Weeks 2-4), and in the last two weeks of the intervention period (Weeks 5-6). p – values are for between-group comparisons using linear mixed models.

**Table 2.**
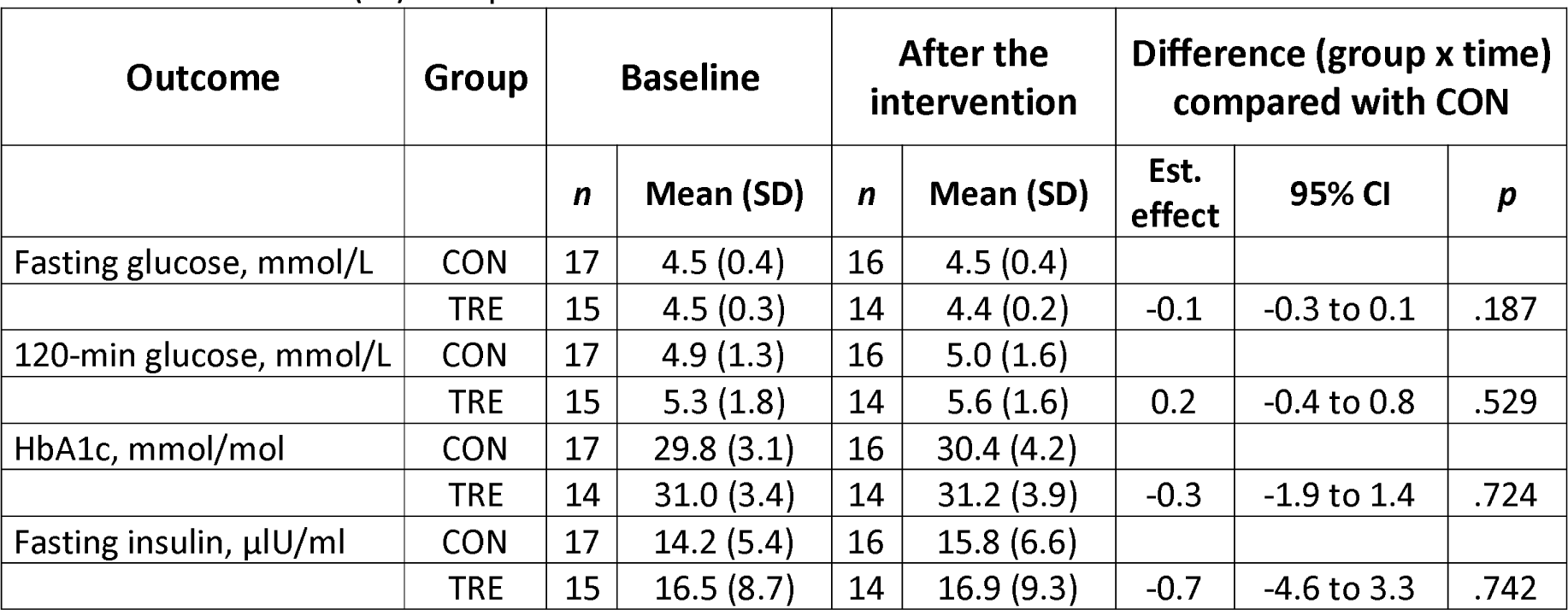
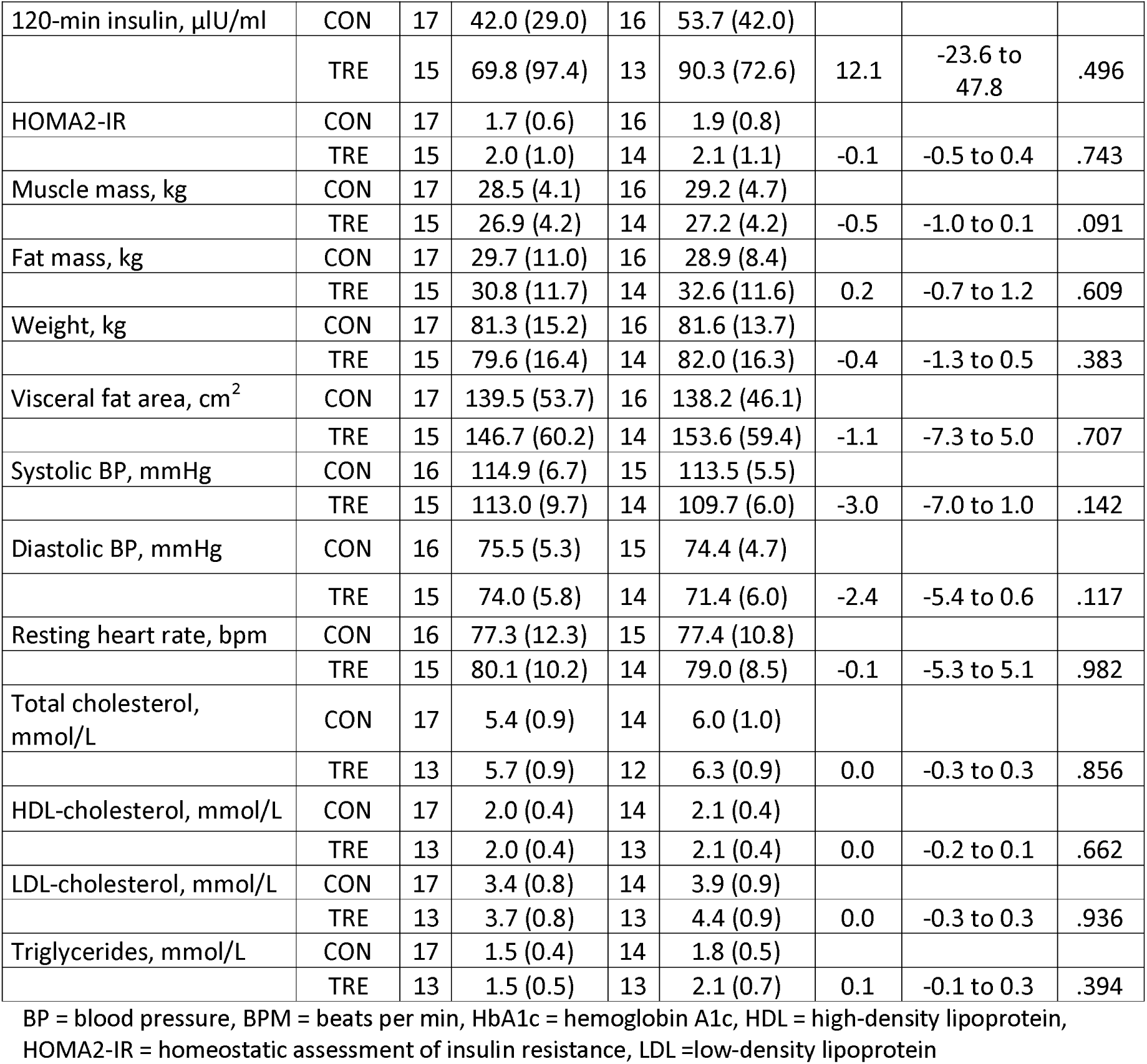
Intention-to-treat analyses of secondary outcomes. Baseline data are reported as mean values with standard deviations (SD) of observed values at baseline, and post-intervention for n participants in each group. Results from linear mixed model analysis presents estimated effect, which represents the difference in mean in the time-restricted eating group (TRE) compared with the control group (CON), with corresponding 95% confidence interval (CI) and p-values.

### Time-restricted eating had small effect on dietary intake and no effect on physical activity

TRE had no effect on total energy intake (Supplementary Table 3). Participants in the TRE group consumed 33 g less carbohydrates per day in the last week of the intervention period, compared with CON (p = .040). In the second week of the intervention period, participants in the TRE group consumed 34 g less sugar compared with CON (p = .005), but there was no difference between groups in the last week of the intervention. There were no between-group differences in other dietary intake variables (Supplementary Table 3), or in total energy expenditure, or other measures of physical activity throughout the study (Supplementary table 4).

### Adverse events

No adverse events were reported during the study.

## Discussion

This study was the first experimental investigation of the feasibility of a TRE intervention during pregnancy. Our results largely support our main hypothesis that 5 weeks of TRE is feasible during pregnancy, as the participants could adhere to a ≤ 10 h/day eating window on ∼5 days/week throughout the intervention period. The average eating window was just under 10 h/day in the intervention period, representing a 2-h reduction from baseline. Despite this reduced eating window, we failed to detect any beneficial effects of TRE on glycemic control or other metabolic outcomes.

The adherence rate to TRE in our study was 67%, which is somewhat lower than the rates reported in other studies of TRE involving non-pregnant individuals and with a daily eating window of maximum 10 h.^13,16^ We have previously showed that reproductive-aged women with overweight/obesity managed to adhere to an identical TRE intervention on 6.2 days/week (89%) for 7 weeks^13^, while Anton and colleagues reported an adherence rate of 84% to a similar 4-week intervention among overweight, sedentary adults aged 65 years or older.^16^ All these studies had a relatively short intervention period, but Lin et al.^17^ reported 87% adherence to an 8-h TRE protocol over the course of a 12-month study in participants with obesity. The main reason for lower adherence in the present study is likely due to the pregnant state of our participants. In pregnancy, nausea and preference of specific foods (cravings) are common, especially in the first trimester. These factors may affect the ability to consume energy only at specific periods during the day as required for TRE. Indeed, a qualitative study on attitudes towards TRE among people who were pregnant or had recently given birth, reported that some were concerned about the baby’s health, nausea, and hunger.^14^ However, 47% of the participants in that study perceived TRE as safe during pregnancy, but only 24% of them said they would be willing to try a TRE regimen during pregnancy.^14^

We report no adverse effects of adhering to a 10-h TRE window for 5 weeks and neither do previous studies.^11,13,16-20^ The most commonly reported adverse effects of TRE are nausea, headaches, dizziness, diarrhea, and dry mouth. However, these effects will either diminish over time, or are resolved by increasing water intake.^11,16,18^. Despite that a 10-h TRE window was found to be feasible in the current study, including participants earlier in pregnancy could yield different results. The mean gestational age in the intervention group in our study was 18.6 weeks (ranging from 12 to 30 weeks) at baseline, whereas nausea is most common in early pregnancy.^21^ The on-going BEFORE THE BEGINNING trial will determine the feasibility of TRE also in early pregnancy.^22^

There was no effect of TRE on any of the measured cardiometabolic outcomes in our study. A reason for this could be that 5 weeks may be insufficient to induce any significant glycemic changes in pregnant individuals.^23^ However, in non-pregnant individuals, TRE interventions do improve glycemic control after interventions of similar duration.^13,18,24^ In one study, restricting the window for energy intake to between 08:00 and 16:00 h improved skeletal muscle insulin sensitivity among healthy males after 2 weeks.^24^ Similar results were also seen in a crossover trial involving men with prediabetes, in which 5 weeks of TRE with a 6-h eating window early in the day reduced the concentrations of insulin both in the fasting state and during a 3-h OGTT.^18^ In both these studies, the window of energy intake was substantially shorter than in the present study, which may explain the different findings. In our previous study involving reproductive-aged women with overweight/obesity, we showed lower nocturnal glucose after 7 weeks of TRE with a 10-h window for energy intake.^13^ In contrast to our previous trial, the participants in the present study did not reduce their energy intake during the intervention period.

Concurrent with no change in energy intake or expenditure, 5 weeks of TRE did not affect body weight. Weight loss is often the goal of TRE interventions^11,25-27^, with improvements in glycemic measures frequently occurring after weight loss.^13^ Even if most TRE regimens allow unrestricted intake of energy within the stipulated eating window, people typically reduce their daily energy intake unintentionally, which likely underpins most of the cardiometabolic benefits of TRE.^28^ The weight loss observed after TRE interventions makes it difficult to determine whether there is a weight loss-independent effect of TRE. However, in the study by Sutton and colleagues, in which the participants were provided with standardized meals to maintain energy balance and weight, they observed several improvements in glycemic outcomes.^18^ Conversely, others have reported reductions in body weight after a TRE intervention without concomitant improvements in measures of glycemic control.^27^ Additionally, we found no change in blood pressure or resting heart rate after the intervention, which is in concordance with findings in similar trials.^11,13,27^ Conversely, some trials have reported reduced blood pressure following TRE interventions.^18,29-31^

We placed no restrictions on the timing of the eating window, but we recommended that the participants started their daily energy intake by 09:00 hrs. On average, participants commenced their daily eating window at 08:43 h and ended it at 18:50 h during the intervention. Placing the eating window early in the day has been shown to improve glycemic outcomes in animals and humans^18,32,33^, and it is suggested to be beneficial for glucose metabolism and insulin sensitivity to synchronize meal timing with the circadian rhythm. Implementing an eating schedule synchronized with the body’s natural activity-rest cycles has been shown to lower blood glucose and insulin concentrations.^33,34^ In pregnant individuals, it was recently shown that consuming > 50% of total daily energy intake between 19:00 and 07:00 h was associated with less desirable glycemic outcomes, including increased fasting glucose and higher 24-h glucose levels.^35^ A previous study by the same research group also indicated that increased maternal night-fasting intervals were associated with decreased fasting glucose in the late-second trimester of pregnancy.^36^Furthermore, in non-pregnant individuals with overweight or obesity, early TRE has superior effects compared with later TRE in improving glycemic control.^37^

The participants in our study had increased risk of developing GDM, but none had signs of abnormal glucose metabolism at baseline. A more pronounced effect of TRE on glycemic outcomes would be more likely if the participants’ glucose metabolism was compromised.^23^ As such, TRE has demonstrated improved cardiometabolic outcomes in people with impaired glucose metabolism, including individuals with the metabolic syndrome, pre-diabetes, or type 2 diabetes.^18,19,31,38,39^

There are several limitations to this study and the interpretation of results. The participants who volunteered for our study were probably less bothered by nausea than the general population of pregnant people with increased risk of GDM, and our intervention period may have been too short to elicit glycemic or cardiometabolic changes. There is also a possibility that the duration of the daily eating window in our trial was too long, or it was set too late in the day to impact glycemic control. Considering the unique physiological state of pregnancy, our results are difficult to compare with studies in non-pregnant populations. Pregnant individuals require special considerations and adjustments, and the metabolic response in pregnant people could differ significantly from that of non-pregnant individuals.

### Conclusion

To the best of our knowledge, the present study is the first trial to determine the feasibility of TRE in pregnancy. While the results of our study suggest that TRE is feasible in the second and third trimesters of pregnancy among individuals at risk of GDM, there were no improvements in glycemic or cardiometabolic outcomes in our subject cohort. Further studies should include a larger number of participants and implement TRE beyond 5 weeks to determine the long-term feasibility and potential health benefits of TRE during pregnancy.

## Methods

### Study design and participants

This parallel-group RCT was carried out at the Norwegian University of Science and Technology (NTNU) and St. Olav’s hospital. The study was approved by the Regional Committees for Medical and Health Research Ethics in Norway (ID 12366) and registered in Clinical Trials (ClinicalTrials.gov, identifier: NCT03803072). All participants provided written informed consent before participating in the study. Participants were recruited through public advertising at St. Olav’s hospital, on social media, and university web pages. We screened for eligibility via telephone before we included participants in the trial. Table 3 shows the inclusion and exclusion criteria. The study period for each participant was 6 weeks, including one week of baseline measurements and 5 weeks of TRE for the intervention group or no intervention for participants in control group. Fasting venous blood sampling, OGTT, estimation of body composition, blood pressure and heart rate measurements were undertaken at the baseline visit and after the 5-week intervention period (Figure 7). After baseline assessments were completed, the participants were randomly allocated (1:1) to either 5 weeks of TRE or usual care (CON). All participants were fitted with continuous glucose monitors (CGMs) and physical activity monitors and received instructions on how to self-report their daily eating window in the study handbook. Dietary intake and physical activity were measured for 7 days in the baseline week, in the second week of the intervention, and in the last week of the intervention (Figure 7).

**Figure 7.**
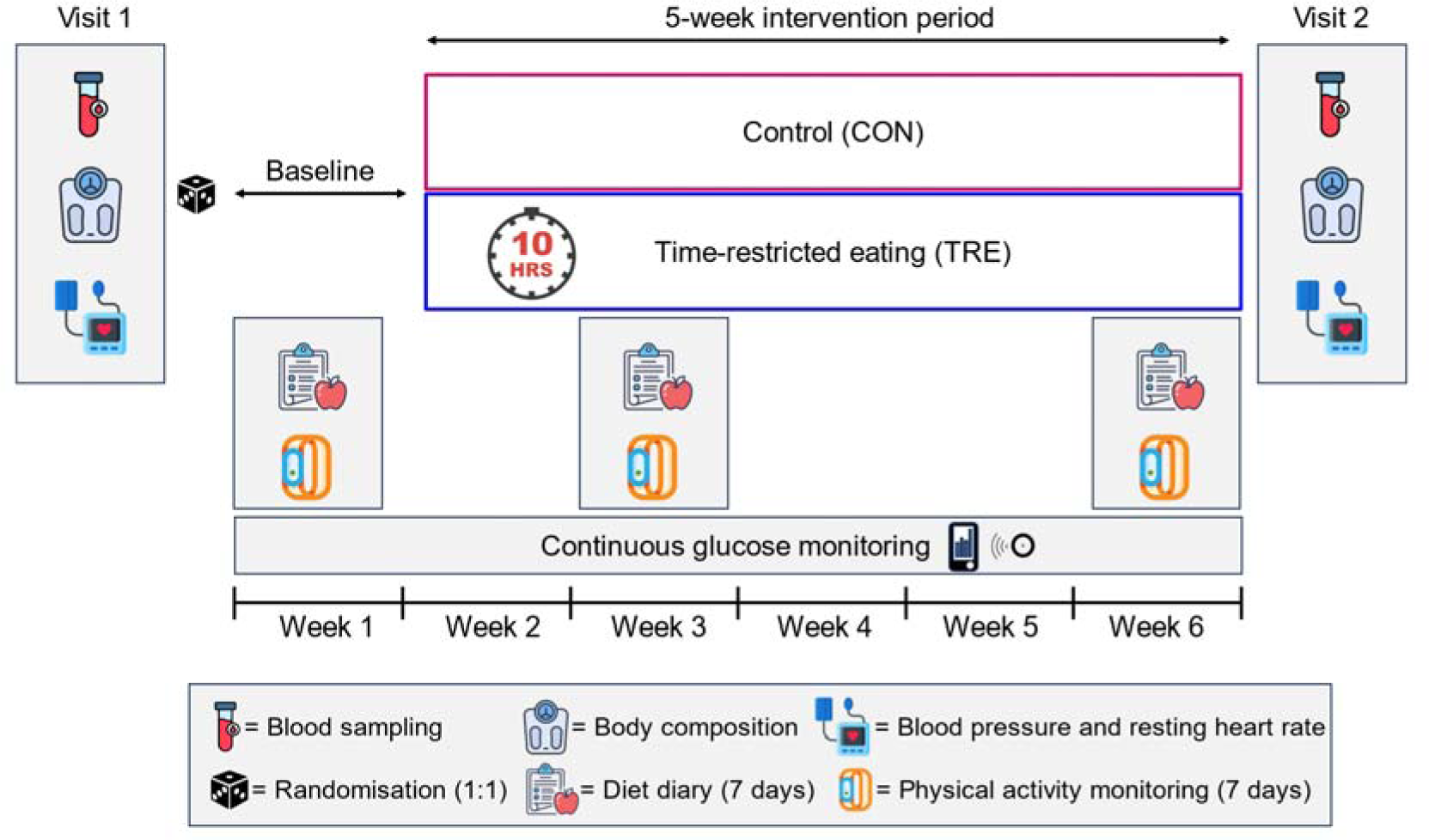
Study design. Participants visited the laboratory for assessments at baseline before randomization, and after the intervention. These visits included fasting blood sampling, an oral glucose tolerance test, estimation of body composition, blood pressure and heart rate measurements. The participants wore continuous glucose monitors throughout the study and physical activity monitors in the baseline week, week 3, and week 6. They registered the time points for first and last energy intake each day throughout the study and reported dietary intake in the baseline week, week 3, and week 6. baseline week of the study.

**Table 3.** Inclusion and exclusion criteria.

| Inclusion criteria | Exclusion criteria |
| --- | --- |
| • Age $\geq 18$ years<br>• Pregnant with a singleton foetus in gestational week 12-30<br>• Understands written and spoken Norwegian or English<br>• At least one risk factor for gestational diabetes (GDM) as defined by Norwegian guidelines for GDM screening<sup>15</sup>:<br>○ Pre-pregnancy body mass index $\geq 25$ kg/m<sup>2</sup><br>○ First birth at the age of $\geq 25$ years<br>○ Previous GDM<br>○ Previous delivery of newborn $\geq 4.5$ kg<br>○ First degree relative with diabetes mellitus<br>○ Asian or African ethnicity | • Habitual daily eating window $\leq 12$ h<br>• Previously diagnosed with diabetes type 1 or 2 |

### Randomization and blinding

The participants undertook baseline assessments before being randomly allocated (1:1) to either TRE or CON, after stratification for BMI < 27 kg/m^2^ or ≥ 27 kg/m^2^. The first or the last author performed the randomization using a random number generator (WebCRF, The Unit for Applied Clinical Research, NTNU, Trondheim). The randomization sequenced was concealed in the WebCRF until interventions were assigned. Neither participants nor study personnel were blinded.

### Intervention

All participants continued with their habitual eating habits during the baseline week. The participants allocated to TRE were instructed to limit their time-window for energy intake to maximum 10 h/day for 5 weeks. We advised the participants to start their eating window no later than 9:00 h and to consume their last energy intake no later than 19:00 h. They could freely consume energy-free beverages such as black coffee, tea, and diet soda outside their daily eating window. We gave no advice regarding the amount of energy or types of foods to be consumed. Participants allocated to CON were instructed to continue their habitual eating pattern for the entire 6-week study period. All participants received a booklet about healthy lifestyle habits in pregnancy.

### Primary outcomes

The primary outcome measure in this study was the feasibility of TRE during pregnancy. Adherence to TRE was self-reported in a study handbook, in which the participants recorded the time points of their first and last energy intake every day throughout the whole study period. We calculated adherence as the average duration of the daily eating window throughout the 5-week study period, as well as the average number of days per week the participants adhered to the ≤ 10-h eating window.

### Secondary outcomes

#### Blood analyses

We sampled venous blood at baseline and after 6 weeks. The participants attended the laboratory after a 10-h overnight fast. After the fasting blood sample was obtained, the participants underwent a 120-min OGTT in which they ingested 75 g glucose dissolved in 250 mL water (GlucosePro, Norges Naturmedisinsentral AS). A second venous blood sample was obtained 120 min after the ingestion of the glucose solution. Glycemic outcomes include fasting plasma glucose, 120-min plasma glucose after the OGTT, HbA1c, fasting serum insulin, and 120-min serum insulin after the OGTT. Additionally, fasting blood total cholesterol, triglycerides, high-density lipoprotein (HDL) and low-density lipoprotein (LDL) were measured. EDTA tubes were centrifuged immediately after sampling at 2220G and 4°C for 10 min. Serum tubes rested upright for 30 min before being centrifuged at 2220G and 20°C for 10 min. All analyses apart from insulin concentrations were carried out at the laboratory at St. Olav’s hospital. Additional aliquots of plasma, serum and full blood were stored at -80C for later analysis. We analyzed fasting and 120-min serum insulin using enzyme-linked immunosorbent assay (ELISA, IBL-International, Hamburg, Germany). These analyses were carried out per manufacturer’s instructions using a DS2 ELISA processing system (Dynex Technologies, Virginia, USA) at the Department of Circulation and Medical Imaging, NTNU. Using fasting blood glucose levels and fasting insulin levels, we calculated insulin resistance (HOMA2-IR) using the online HOMA2 calculator (https://www.rdm.ox.ac.uk/about/our-clinical-facilities-and-mrc-units/DTU/software/homa).

#### Continuous glucose monitoring

The participants wore CGMs (FreeStyle Libre 1, Abbott Diabetes Care, Norway) throughout the entire 6-week study period. They were fitted with CGM sensors at baseline and instructed to do minimum four scans evenly spaced out throughout each day of the study. We gave out replacement sensors at baseline to cover the whole 6-weeks study duration. We covered the screen of the CGM monitor to avoid that the participants got aware of their glucose levels. Raw CGM data were processed using Microsoft Excel and divided into a 1-week baseline period (week 1), a 3-week mid-study period (weeks 2-4) and 2-week end-period (weeks 5-6). We used the Glyculator 3.0 calculator (https://glyculator.btm.umed.pl) to impute missing glucose measurements due to infrequent scans and to estimate glucose AUC for 24 h, daytime (06:00 – 00:00) and night-time (00:01 – 05:59).

#### Body composition, blood pressure, and resting heart rate

We estimated the participants’ body composition using a bioelectrical impedance scale (InBody770, Biospace CO, Ltd, Seoul, Korea) after a ≥ 10 h overnight fast. The participants wore light clothing and no shoes during these tests. Parameters used to estimate body composition include height, weight, BMI, fat mass, muscle mass and visceral fat area. Blood pressure and heart rate were measured using an automated blood pressure device (Welch Allyn, Germany). We obtained three measurements with 1-min intervals and report the average of these three measurements.

#### Physical activity and diet

We fitted the participants with physical activity monitors (BodyMedia Sensewear Armband, Pittsburgh, PA) during the baseline testing. These monitors were worn during the baseline week, the 3^rd^ study week, and the 6^th^ study week to estimate average weekly physical activity and energy expenditure. Raw physical activity data were processed using Microsoft Excel and divided into baseline (week 1), mid-study (weeks 2-4), and end-period (weeks 5-6). In the same periods, the participants recorded their dietary intake for 7 days using an online food diary (kostholdsplanleggeren.no). Raw dietary data were processed using Microsoft Excel. In the same weeks, participants recorded subjective feelings of hunger, fullness, satiety, and desire to eat in the mornings and evenings on 10-cm visual analogue scales printed in their study handbooks.

### Sample size

We did not perform a formal power analysis for this trial. Generally, a sample size between 24 and 50 is recommended to estimate effect size and standard deviation in feasibility studies ^40-42^. We aimed to include 32 participants to ensure that we had a minimum of 24 (12 in each group) with measurements of glucose tolerance and insulin sensitivity at two time points, accounting for an expected drop-out of 20%.

### Statistical analysis

The adherence data are reported as descriptive statistics. We used linear mixed models (LMM) to compare adherence in TRE with CON. In the LMM models, we included time and the interaction between time and group as fixed factors, and participants as random factor.^43^ We report the estimated mean change in the TRE group, with corresponding 95% confidence intervals (CI) and p-values, compared with CON. We also used linear mixed models to compare secondary outcome measures between TRE and CON. All randomized participants were included in the intention-to-treat analysis, regardless of adherence. We excluded data with less than 4 days of valid CGM measurements or physical activity data in a period, as well as less than at least two weekdays and one weekend day of nutritional intake. However, if the participant had sufficient data in other time periods, then these data were included in the analyses. Statistical analyses were performed in IBM SPSS Statistics 27. We consider p-values < .05 as statistically significant and have not performed any adjustments for multiple comparisons due to the exploratory nature of our research questions.

## Supporting information

CONSORT checklist

## Data availability

Data reported in this paper will be shared by the corresponding author upon reasonable request.

## Author contributions

TM, KÅS, and JAH conceived the study and analysis plan. HMS, KLH, TMG, and MKJ collected the data. HMS, MAJS, KLH and TM analyzed the data. All authors contributed to the interpretation of the data. HMS drafted the manuscript. All authors critically reviewed the manuscript for intellectual content and gave their final approval of the version to be published.

## Acknowledgements

This work was supported by the Norwegian University of Science and Technology (NTNU), by a Novo Nordisk Foundation Challenge Grant to JAH (NNF14OC0011493), an EFSD and Novo Nodisk Foundation Future Leaders Award Programme grant to TM (NNF19SA058975), and by The Liaison Committee for Education, Research, and Innovation in Central Norway (2020/39645). The funding bodies had no role in the design, data collection, or interpretation of results. We wish to thank the Unit for Applied Clinical Research at NTNU for providing the internet-based randomization, Elisabeth Eide Axe and Guro Rosvold for assistance with blood sampling and data collection, and the Department of Clinical Chemistry at St. Olavs Hospital for biochemical analyses of blood samples. Finally, we want to thank all the participants in the trial.

## Materials & Correspondence

Correspondence and material requests should be addressed to Trine Moholdt

