## Supplementary Tables for "Feasibility of time-restricted eating during pregnancy and effect on glycemic control in people with increased risk of gestational diabetes – a randomized controlled trial"

**Supplementary Table 1.** Ratings of hunger and appetite on visual analogue scales in the morning and in the evening. Data are observed values with standard deviation (SD) at baseline (Week 1), the second week of the intervention (Week 3), and in the last week of the intervention (Week 6). Between-group differences are estimated using linear mixed models and reported as estimated (est.) effects with 95% confidence intervals (CI) and p-values.

|  |  | **Week 1** | **Week 3** | **Week 6** | **Between-group difference**  **Week 3** | | | **Between-group difference**  **Week 6** | | |
| --- | --- | --- | --- | --- | --- | --- | --- | --- | --- | --- |
| **Outcome** | **Group** | **Mean (SD)** | **Mean (SD)** | **Mean (SD)** | **Est. effect** | **95% CI** | **p** | **Est. effect** | **95% CI** | **p** |
| **Morning ratings** | | | | | | | | | | |
| Hunger | CON | 4.6 (2.3) | 4.6 (1.9) | 4.3 (2.2) |  |  |  |  |  |  |
|  | TRE | 5.5 (1.6) | 5.6 (1.9) | 5.6 (1.9) | 0.1 | -0.5 to 0.7 | .689 | 0.5 | -0.1 to 1.2 | .078 |
| Fullness | CON | 2.8 (2.1) | 3.0 (1.9) | 3.0 (2.0) |  |  |  |  |  |  |
|  | TRE | 2.6 (1.7) | 2.2 (1.4) | 2.3 (1.6) | -0.4 | -0.9 to 0.1 | .096 | -0.4 | -0.9 to 0.1 | .156 |
| Satiety | CON | 2.5 (2.0) | 2.9 (1.9) | 2.9 (2.0) |  |  |  |  |  |  |
|  | TRE | 2.5 (1.7) | 2.3 (1.5) | 2.3 (1.5) | -0.4 | -0.9 to 0.1 | .075 | -0.4 | -0.9 to 0.1 | .127 |
| Desire to eat | CON | 5.3 (2.4) | 5.2 (2.0) | 5.0 (2.1) |  |  |  |  |  |  |
|  | TRE | 6.1 (1.8) | 5.9 (1.9) | 6.3 (2.0) | 0.1 | -0.5 to 0.7 | .270 | 0.6 | -0.02 to 1.2 | .056 |
| Prospective intake | CON | 4.9 (1.9) | 4.8 (1.7) | 4.8 (1.9) |  |  |  |  |  |  |
|  | TRE | 5.6 (1.8) | 5.7 (1.7) | 5.8 (2.0) | 0.2 | -0.2 to 0.8 | .422 | 0.4 | -0.1 to 0.9 | .144 |
| **Evening ratings** | | | | | | | | | | |
| Hunger | CON | 2.0 (1.9) | 2.1 (1.7) | 2.2 (1.7) |  |  |  |  |  |  |
|  | TRE | 2.3 (1.8) | 3.6 (2.1) | 3.3 (2.2) | 1.5 | 0.9 to 2.1 | <.001 | 0.9 | 0.3 to 1.5 | .002 |
| Fullness | CON | 5.7 (2.1) | 5.5 (2.1) | 5.8 (1.9) |  |  |  |  |  |  |
|  | TRE | 5.2 (1.8) | 4.5 (2.3) | 4.9 (2.2) | -0.6 | -1.3 to 0.04 | .066 | -0.4 | -1.1 to 0.3 | .231 |
| Satiety | CON | 5.4 (2.0) | 5.5 (2.2) | 5.6 (1.9) |  |  |  |  |  |  |
|  | TRE | 5.3 (1.8) | 4.4 (2.3) | 4.8 (2.1) | -0.9 | -1.6 to -0.3 | .007 | -0.7 | -1.4 to -0.04 | .037 |
| Desire to eat | CON | 2.4 (2.0) | 2.4 (1.9) | 2.6 (1.7) |  |  |  |  |  |  |
|  | TRE | 2.8 (2.0) | 4.0 (2.4) | 3.5 (2.2) | 1.5 | 0.8 to 2.1 | <.001 | 0.6 | -0.02 to 1.3 | .058 |
| Prospective intake | CON | 2.4 (2.0) | 2.5 (1.8) | 2.8 (1.9) |  |  |  |  |  |  |
|  | TRE | 2.7 (1.8) | 3.9 (2.3) | 3.6 (2.2) | 1.1 | 0.5 to 1.7 | <.001 | 0.5 | -0.08 to 1.2 | .088 |

**Supplementary Table 2.** Glucose area under the curve (AUC) for 24 hours (24-h), during the daytime (Day) and night-time (Night), estimated from continuous glucose monitors. Data are observed values with standard deviation (SD) at baseline, the second week of the intervention (Week 3), and in the last week of the intervention (Week 6). Between-group differences are estimated using linear mixed models and reported as estimated (est.) effects with 95% confidence intervals (CI) and p-values.

|  |  | **Week 1** | **Weeks**  **2-4** | **Weeks 5-6** | **Between-group difference**  **Week 3** | | | **Between-group difference**  **Week 6** | | |
| --- | --- | --- | --- | --- | --- | --- | --- | --- | --- | --- |
| **Outcome** | **Group** | **Mean (SD)** | **Mean (SD)** | **Mean (SD)** | **Est. effect** | **95% CI** | ***p*** | **Est. effect** | **95% CI** | ***p*** |
| 24-h AUC, mmol/L | CON | 4.5 (0.4) | 4.5 (0.4) | 4.4 (0.5) |  |  |  |  |  |  |
|  | TRE | 4.5 (0.9) | 4.6 (0.2) | 4.8 (0.4) | 0.01 | -0.2 to 0.3 | .953 | 0.24 | -0.01 to 0.5 | .059 |
| Day AUC, mmol/L | CON | 4.7 (0.4) | 4.6 (0.4) | 4.5 (0.5) |  |  |  |  |  |  |
|  | TRE | 4.6 (0.9) | 4.8 (0.3) | 5.0 (0.4) | 0.02 | -0.2 to 0.3 | .855 | 0.24 | -0.02 to 0.5 | .074 |
| Night AUC, mmol/L | CON | 4.0 (0.5) | 4.1 (0.3) | 4.0 (0.5) |  |  |  |  |  |  |
|  | TRE | 3.9 (0.8) | 4.1 (0.2) | 4.1 (0.4) | -0.07 | -0.3 to 0.2 | .624 | 0.11 | -0.2 to 0.4 | .442 |

**Supplementary Table 3.** Self-reported daily dietary intake. Data are observed values with standard deviation (SD) at baseline (Week 1), the second week of the intervention (Week 3), and in the last week of the intervention (Week 6). Between-group differences are estimated using linear mixed models and reported as estimated (est.) effects with 95% confidence intervals (CI) and p-values.

|  |  | **Week 1** | **Week 3** | **Week 6** | **Between-group difference**  **Week 3** | | | **Between-group difference**  **Week 6** | | |
| --- | --- | --- | --- | --- | --- | --- | --- | --- | --- | --- |
| **Outcome** | **Group** | **Mean (SD)** | **Mean (SD)** | **Mean (SD)** | **Est. effect** | **95% CI** | ***p*** | **Est. effect** | **95% CI** | ***p*** |
| Total energy intake, kJ | CON | 9285 (2492) | 9281 (3202) | 8855 (2415) |  |  |  |  |  |  |
|  | TRE | 9012 (1293) | 8884 (2280) | 8920 (2223) | -15 | -970 to 940 | .975 | -184 | -1139 to 772 | .701 |
| Fat, g | CON | 91 (30) | 89 (40) | 80 (29) |  |  |  |  |  |  |
|  | TRE | 85 (19) | 86 (32) | 90 (32) | 5 | -10 to 21 | .496 | 9 | -7 to 24 | .264 |
| Saturated fat, g | CON | 37 (16) | 37 (19) | 31 (13) |  |  |  |  |  |  |
|  | TRE | 34 (9) | 33 (15) | 34 (15) | -4 | -10 to 3 | .256 | 2 | -4 to 8 | .548 |
| Cis-mono-unsaturated fat, g | CON | 33 (12) | 32 (17) | 29 (13) |  |  |  |  |  |  |
|  | TRE | 30 (6) | 32 (16) | 34 (14) | 5 | -4 to 13 | .276 | 4 | -4 to 13 | .341 |
| Cis-poly-unsaturated fat, g | CON | 12 (4) | 11 (5) | 12 (7) |  |  |  |  |  |  |
|  | TRE | 13 (4) | 13 (6) | 14 (6) | 3 | -0.3 to 5 | .081 | 1 | -2 to 4 | .483 |
| Carbohydrates, g | CON | 245 (74) | 255 (100) | 254 (79) |  |  |  |  |  |  |
|  | TRE | 241 (34) | 240 (77) | 227 (71) | -14 | -46 to 18 | .388 | -33 | -65 to -2 | .040 |
| Sugar, g | CON | 40 (26) | 66 (58) | 42 (27) |  |  |  |  |  |  |
|  | TRE | 53 (50) | 37 (35) | 31 (28) | -34 | -58 to -11 | .005 | -16 | -40 to 9 | .198 |
| Fiber, g | CON | 24 (6) | 23 (10) | 24 (9) |  |  |  |  |  |  |
|  | TRE | 25 (5) | 22 (7) | 24 (8) | -2 | -5 to 1 | .187 | -2 | -5 to 1 | .277 |
| Protein, g | CON | 93 (17) | 92 (30) | 85 (28) |  |  |  |  |  |  |
|  | TRE | 92 (11) | 86 (28) | 91 (26) | -3 | -12 to 6 | .513 | 1 | -8 to 11 | .792 |

kJ = kilojoules, TEI = Total energy intake.

**Supplementary Table 4.** Daily physical activity, estimated from activity armbands. Data are observed values with standard deviation (SD) at baseline (Week 1), the second week of the intervention (Week 3), and in the last week of the intervention (Week 6). Between-group differences are estimated using linear mixed models and reported as estimated (est.) effects with 95% confidence intervals (CI) and p-values.

|  |  | **Week 1** | **Week 3** | **Week 6** | **Between-group difference**  **Week 3** | | | **Between-group difference**  **Week 6** | | |
| --- | --- | --- | --- | --- | --- | --- | --- | --- | --- | --- |
| **Outcome** | **Group** | **Mean (SD)** | **Mean (SD)** | **Mean (SD)** | **Est. effect** | **95% CI** | ***p*** | **Est. effect** | **95% CI** | ***p*** |
| Physical activity level, METs | CON | 1.2 (0.2) | 1.3 (0.2) | 1.3 (0.1) |  |  |  |  |  |  |
|  | TRE | 1.3 (0.2) | 1.3 (0.3) | 1.4 (0.4) | -0.01 | -0.2 to 0.2 | .934 | 0.1 | -0.1 to 0.3 | .188 |
| Daily energy expenditure, kJ | CON | 8888 (1513) | 8967 (1490) | 8498 (1421) |  |  |  |  |  |  |
|  | TRE | 9176 (962) | 9430 (911) | 10245 (602) | 63 | -765 to 890 | .879 | 794 | -68 to 1655 | .070 |
| Sedentary time, min | CON | 1068 (184) | 943 (183) | 960 (243) |  |  |  |  |  |  |
|  | TRE | 920 (209) | 927 (231) | 995 (289) | 16 | -106 to 138 | .789 | -31 | -157 to 96 | .627 |
| Light intensity activity, min | CON | 142 (62) | 183 (141) | 165 (93) |  |  |  |  |  |  |
|  | TRE | 166 (111) | 180 (114) | 197 (86) | 1 | -63 to 64 | .981 | 12 | -54 to 77 | .723 |
| Moderate intensity activity, min | CON | 50 (45) | 52 (48) | 42 (30) |  |  |  |  |  |  |
|  | TRE | 49 (35) | 57 (51) | 75 (66) | 7 | -24 to 37 | .660 | 26 | -5 to 58 | .097 |
| Vigorous intensity activity, min | CON | 4 (9) | 3 (6) | 3 (5) |  |  |  |  |  |  |
|  | TRE | 2 (2) | 4 (7) | 6 (8) | 1 | -4 to 7 | .577 | 4 | -2 to 9 | .149 |

METs = Metabolic equivalent of task, PA = physical activity
